# Psychometric Evaluation of the Snakebite Severity Score (SSS) in a Multinational Randomized Clinical Trial

**DOI:** 10.64898/2026.06.22.26356259

**Authors:** João Vítor Perez de Souza, William Nkenguye, Grayson Chappell, Timothy F. Platts-Mills, João Ricardo Nickenig Vissoci, Charles J Gerardo

## Abstract

**Background:** Snakebite envenomation (SBE) is a World Health Organization recognized neglected tropical disease (NTD) affecting 1.8 million annually. Currently SBE research lacks standardized, patient-centered outcome measures, hindering comparability and clinical relevance. The Snakebite Severity Score (SSS) is a composite endpoint developed to assess symptom severity across multiple body systems. The present study evaluates the psychometric properties of the SSS using data from the BRAVO Phase 2b clinical trial of varespladib-methyl.

**Methods:** A secondary analysis was conducted using data from the BRAVO clinical trial (NCT04996264), a randomized, double-blind, placebo-controlled Phase 2b study evaluating varespladib-methyl. Patients aged ≥5 years with symptomatic SBE were enrolled from emergency departments in India and the US. The SSS and modified versions (6-item and 3-item) were administered at baseline and at multiple follow-up time points: 3, 6, and 9 hours post-envenomation, and on days 2, 3, 7, 14, and 28. Psychometric analyses included descriptive statistics, intraclass correlation coefficients (ICC) for reliability, principal component analysis (PCA) for internal structure, and correlations with patient-reported outcomes (PSFS, PGIC, NPRS) and clinician-rated CGI-I for external validity.

**Results:** Ninety-five participants were analyzed (varespladib: n=45; placebo: n=50). The 6-item SSS demonstrated strong reliability (ICC = 0.8 at Days 7-14) and consistent internal structure across subscores. PCA confirmed multidimensionality, with distinct contributions from local wound, nervous system, hematological, and other subscales. External validity was supported through moderate to strong correlations with PGIC, NPRS, and CGI-I, particularly for applications capturing symptom variation over time (AUC, mean scores). The 6-item SSS captured symptom severity more robustly than the 3-item version.

**Conclusion:** The SSS is a reliable and valid multidimensional composite endpoint for assessing clinical severity in SBE. Applications that integrate symptom change over time demonstrate better external validity and are preferable. Findings support SSS use in clinical research to standardize and improve outcome assessment in SBE.

## INTRODUCTION

Snakebite envenomation (SBE) is a WHO category A neglected tropical disease that affects an estimated 1.8 million people. Of these, an estimated 63,400 -125,000 do not survive and three to four times as many are permanently disabled(1–3). The global disease burden of SBE is likely underestimated given many SBE victims reside in medically and economically impoverished communities. SBE patients often present to emergency departments (EDs) for initial or advanced care. Research and public health interventions to improve SBE outcomes are crucial to improving the current state(4,5).

Novel therapeutics for SBE are in development for SBE(6–10). However, research is limited by significant heterogeneity of the disease presentation, the lack of agreed upon primary outcomes, and limited psychometric evaluation of the existing outcomes used to assess SBE intervention efficacy(11). A commonly used outcome is the Snakebite Severity Score (SSS). The SSS is a multi-dimensional composite endpoint obtained from the assessment of six key symptomatic venom effects common to SBE: pulmonary, cardiovascular, local wound, gastrointestinal, hematologic, and central nervous system. Each venom effect is scored on a Likert-type scale ranging from 0-3 or 0-4, with a total score range from 0 – 20(12). The SSS was developed as a unique tool to address the complexity of clinical presentation in SBE, that varies by location of the body affected and specific snake toxins. The SSS items were not designed to be a measure where all indicators are theoretically measuring the same latent trait. Rather, the SSS is a composite endpoint designed to quantify the severity of the symptoms experienced by SBE patients and to derive a severity score from an aggregation of the subscores(12).

Although it was originally developed for use in research on North American pit vipers envenoming, it is widely used in SBE research independent of snake taxa or region. With guidance from regulatory bodies, the addition of a new subscore that targets renal toxicity in SBE has also been used in place of the gastrointestinal domain (13). Despite refinements, few studies have evaluated the psychometric properties of the SSS as a measure of SBE outcomes related to different snake species and disease presentations(14,15).

The objective of this study is to evaluate the psychometric properties of the SSS and several of its applications. The study assesses the reliability, internal structure, and external validity of the SSS by conducting a secondary analysis for the BRAVO SBE Clinical Trial. The BRAVO SBE clinical trial is a multi-national, randomized, double-blind, placebo-controlled Phase 2b trial investigating oral varespladib, an inhibitor of phospholipase A2, in SBE treatment(16).

## METHODOLOGY

### Study Design

We performed a psychometric evaluation of the SSS and multiple SSS applications (see below) to characterize disease courses at baseline and across different time points between the treatment arms. This is a secondary analysis using patient data from the BRAVO SBE Trial (NCT04996264), a multinational, randomized, double-blind, placebo-controlled, Phase 2b study designed to evaluate the safety, tolerability, and efficacy of a new oral direct toxin inhibitor, varespladib-methyl. This analysis is reported following the American Psychology Association Standards for Psychological and Educational testing(17) and the COSMIN (Consensus-based Standards for the selection of health Measurement Instruments)(18).

### Participants

All patients meeting eligibility criteria were enrolled in the BRAVO SBE Trial through EDs, followed through with their admission, and had additional assessments at 3, 7, 14, and 28 days after envenomation. In the parent study, patients were randomized to receive standard of care, typically antivenom, and either varespladib-methyl or a placebo. Patients were eligible for inclusion if they were ≥ 5 years of age; having a symptomatic SBE occurring within 10 hours of the trial eligibility assessment and an initial SSS score ≥2 in one component and ≥1 in another or ≥3 in at least one component. The gastrointestinal (GI) category was excluded from SSS inclusion scores. Patients were excluded if they were treated with antivenom prior to trial enrollment; pregnant or breast-feeding; allergic to varespladib-methyl or related compounds; taking an anticoagulant medication; or considered unable to comply with trial protocols due to geographic, psychiatric, or other compliance issues. Patients with an upper GI bleed or a concerning clinical history (see study protocols for details) were also excluded. Written informed consent was obtained from all adults (≥18) participants. Written parental or legal guardian assent was obtained for minors (<18). More information is available in the trial protocol(16).

#### Instruments

##### Snakebite Severity score (SSS) (Modified SSS-6 and 3-item version)

The SSS was completed by the treating physician/investigator. Six subscores were measured, namely local wound, pulmonary, cardiovascular, gastrointestinal, hematological, and nervous system effects(12). The SSS range of severity is 0 to 3 or 0 to 4 for each criterion of evaluation; a higher score indicates more severe effects. The total score ranges from 0 to 20 points. The modified SSS used in this study replaces the GI subscore of the SSS with a renal subscore which was added to the SSS framework based on regulatory guidance and expert opinion(16). This subscore used the previously validated KDIGO tool for assessing acute kidney injury which ranges from 0 to 3(19). The modified SSS was administered by treating physicians/investigators at baseline and post-treatment at 4, 6, and 9 hours, and on days 2, 3, 7, 14, and 28. A three-item SSS, which included the local wound, hematologic and nervous system subscores only, was also evaluated considering the relevance of these criterion for the SBE caused by the snake species in this study.

### SSS applications

There are multiple potential ways to apply the SSS to determine disease course. These SSS applications allow comparison of disease course with different intervention strategies and are described below.

#### SSS Area Under the Curve

The Area Under the Curve (AUC) for the 6-item and 3-item modified SSS are calculated using the trapezoidal rule: average the SSS scores between two adjacent timepoints, multiply by the difference in time points in hours, repeat this across all timepoints during the period of interest, and sum the results to obtain the total AUC across a specified range of time. Thus, the AUC at Day 7 is calculated as follows where BL is baseline, *j* indicates measurement timepoint, SSS*_i,j_* refers to the SSS score for subject *i* at timepoint *j*, and t*_i,j_* refers to the assessment time point for subject *i* at time *j*.

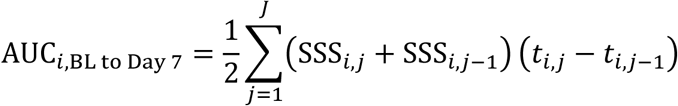

For these calculations, mapped time rather than actual time is used. The maximum scores for the 6-item AUC over 7 day is 3360, which is the product of the maximum SSS score of 20 and 168 hours.

#### Mean SSS over time

The Mean SSS over time is an SSS-derived metrics (3- and 6-Item SSS) that offers alternative ways to assess variations in patient outcomes. These metrics include: Mean SSS between day 3 and day 7 (provides insight into the mid-term trajectory of symptom changes); Mean SSS between day 7 and day 14 (captures longer-term symptom variation and trends over the second week; SSS at day 7 (a snapshot of symptom severity at a key mid-point, offering a critical marker for treatment response).

#### SSS sum of difference

The SSS differences were calculated by subtracting each follow-up time point score from the baseline score (e.g., SSS at baseline - SSS at Hour 3; SSS at baseline – SSS at hour 6, extending through day 14) and summing the differences create the SSS sum of differences. This calculation was applied to both the 3-item SSS (local wound, hematologic, nervous) and the 6-item SSS (all subscores except GI). This approach was informed by analogous methods used in analgesic trials, specifically adaptations of the sum of pain intensity difference (SPID), which summarizes changes in pain over time.(20) It is important to acknowledge that some of the approaches to pain difference from baseline use a time-weight calculation that was not incorporated here.

#### Difference between baseline and 7 or 14 days

The SSS differences between baseline and 7 or 14 days were calculated by subtracting each follow-up time point score (7 and 14 days), separately, from the baseline score (e.g., SSS at baseline - SSS at 7 days; SSS at baseline - SSS at 14 days). This calculation was applied to both the 3-item SSS (local wound, hematologic, nervous) and the 6-item SSS (all items except GI).

##### Patient-Specific Functional Scale (PSFS)

The PSFS is a verbally administered three-item instrument. It evaluates a health condition’s impact on the ability to perform activities chosen by the patient(21). The patient is asked to identify “three activities that you are unable to do or are having difficulty with as a result of your snakebite.” The patient rates each item on an 11-point ordinal scale ranging from 0 (“unable to perform activity”) to 10 (“able to perform activity at the same level as before the injury or problem”). The patient re-rates the same three activities at each follow-up assessment. In this manner, the PSFS collects patient-reported outcomes over time. An average of the three activity scores was used for calculations. This instrument was first administered on day 1, with values recorded at day 7 and day 14

##### Patient Global Impression of Change (PGIC)

PGIC is a widely used research tool to define clinically meaningful improvement in orthopedics, pain, and other diseases(22). This two-item assessment tool uses separate ordinal scales to assess change since the start of treatment. In this study, we only used the first item, a 7-item Likert-type scale anchored at 5 (“moderately better, and a slight but noticeable change”). Patients with scores of ≥5 are considered to have perceived clinically important improvement. Whereas scores <5 have either no change or a clinically unimportant improvement. This instrument was first administered on day 7, with values recorded at day 14.

##### Numeric Pain Rating Scale (NPRS)

The NPRS is an 11-point scale for patient self-reporting pain with scores ranging from 0 (no pain) to 10 (worst possible pain)(23). This instrument was first administered on day 1, with values recorded at day 7 and day 14.

##### Clinical Global Impression - Improvement (CGI-I)

The CGI-I defines clinically meaningful improvement from the perspective of the treating physician. This one-item assessment tool also uses separate ordinal scales to assess change since beginning treatment(24). It is a 7-item Likert-type scale anchored at 4 (“no change from baseline”). Scores <4 indicate clinical improvement and >4 indicate worsening.

##### Ethical Statement

Data used in this analysis was from the BRAVO clinical trial. The trial was registered on Clinical trials.gov, NCT#04996264 and Clinical Trials Registry-India, 2021/07/045079 000062. Prior to performance of any study-related procedures, all patients, or legally authorized representatives, provided written informed consent by signing the ICF after being presented trial information, including procedures, other therapies available, rights, HIPAA compliance, and risks and benefits. Parents of pediatric patients also consent to parental assent.

##### Data collection

In the parent study, following screening and enrollment, each participant provided demographic information, past medical and medication history, and history of the SBE. In addition, a complete physical examination was performed. Data about the initial hospital encounter, including the date and time of arrival, the maximal extent of swelling, medication administration, laboratory test results, and adverse events were recorded by study personnel at the time of care. Formal study assessments were performed at baseline and at multiple follow-up time points: 3, 6, and 9 hours post-envenomation, as well as on days 2, 3, 7, 14, and 28 after the SBE(13).

### Data analysis

All SSS data was described using summary statistics, including mean and standard deviation (SD), median and interquartile range (IQR), and the minimum and maximum values.

#### Psychometric properties

Evidence of reliability was determined by assessing the temporal stability of the SSS using an intraclass correlation (ICC) metric. ICC values between 0.5 and 0.7 were considered moderate and above 0.7 as good reliability(25). Temporal stability was assessed between days 7 and 14, when patients are deemed to be more clinically stable. Evidence of validity related to internal structure was assessed via analysis of correlation and principal component analysis(13). Evidence of validity of SSS related to other instruments was assessed through the correlation of the SSS and the alternative SSS applications with the additional SBE outcomes: CGI-I, NPRS, PGIC, and PSFS.

## RESULTS

Baseline demographic and clinical characteristics of participants in the varespladib (n=45) and placebo (n=50) groups are depicted in Table 1. Data is reproduced from the demographics reported on the parent trial(13). Both groups were similar in age distribution, sex, and geographic location, with most participants being adults aged 18 years or older (N=85, 89%) and enrolled in India (N=62, 65%). The most common snake types were Russell’s viper (N =29, 30%) and Copperhead (N =18, 19%), and average baseline severity scores were slightly higher in the varespladib group than the placebo group 5.9 (SD, 1.9) vs 5.3 (2.2), respectively.

**Table 1:**
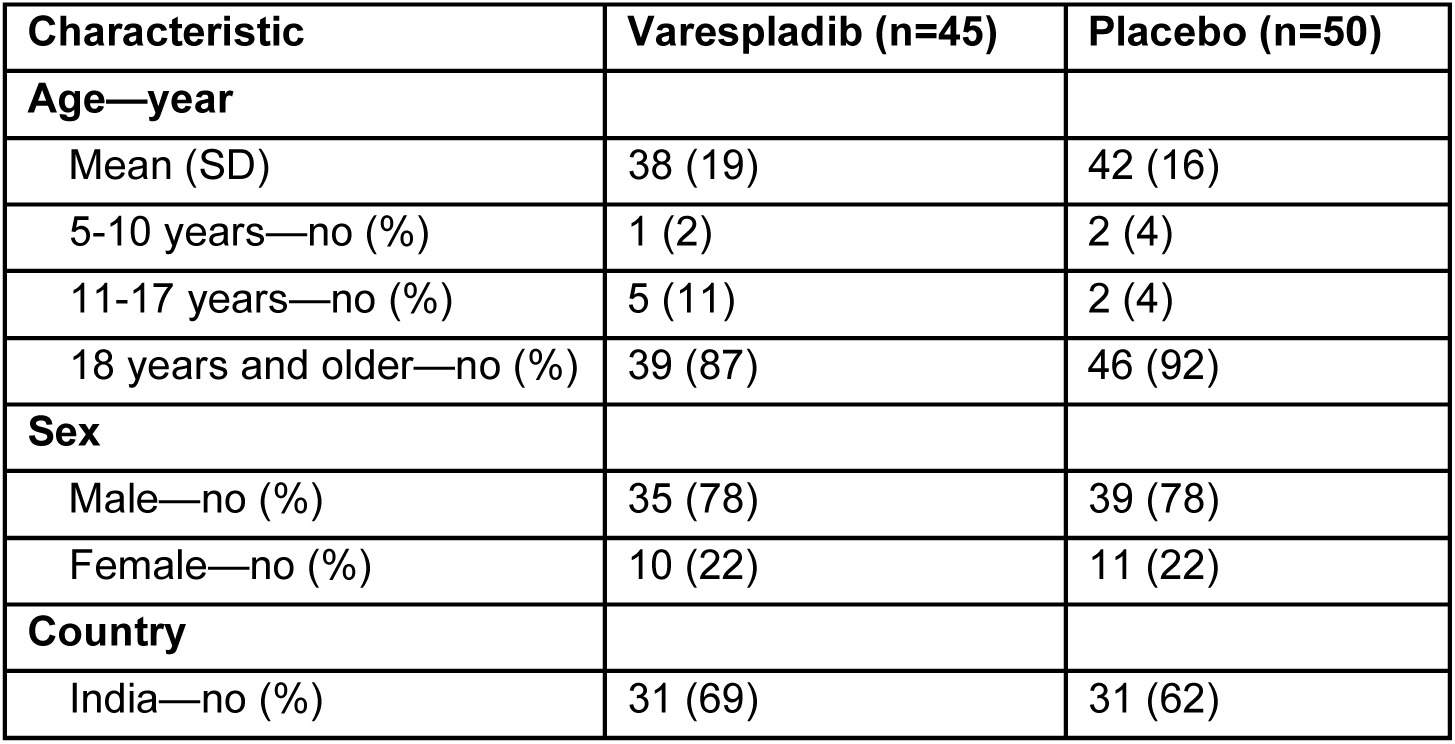
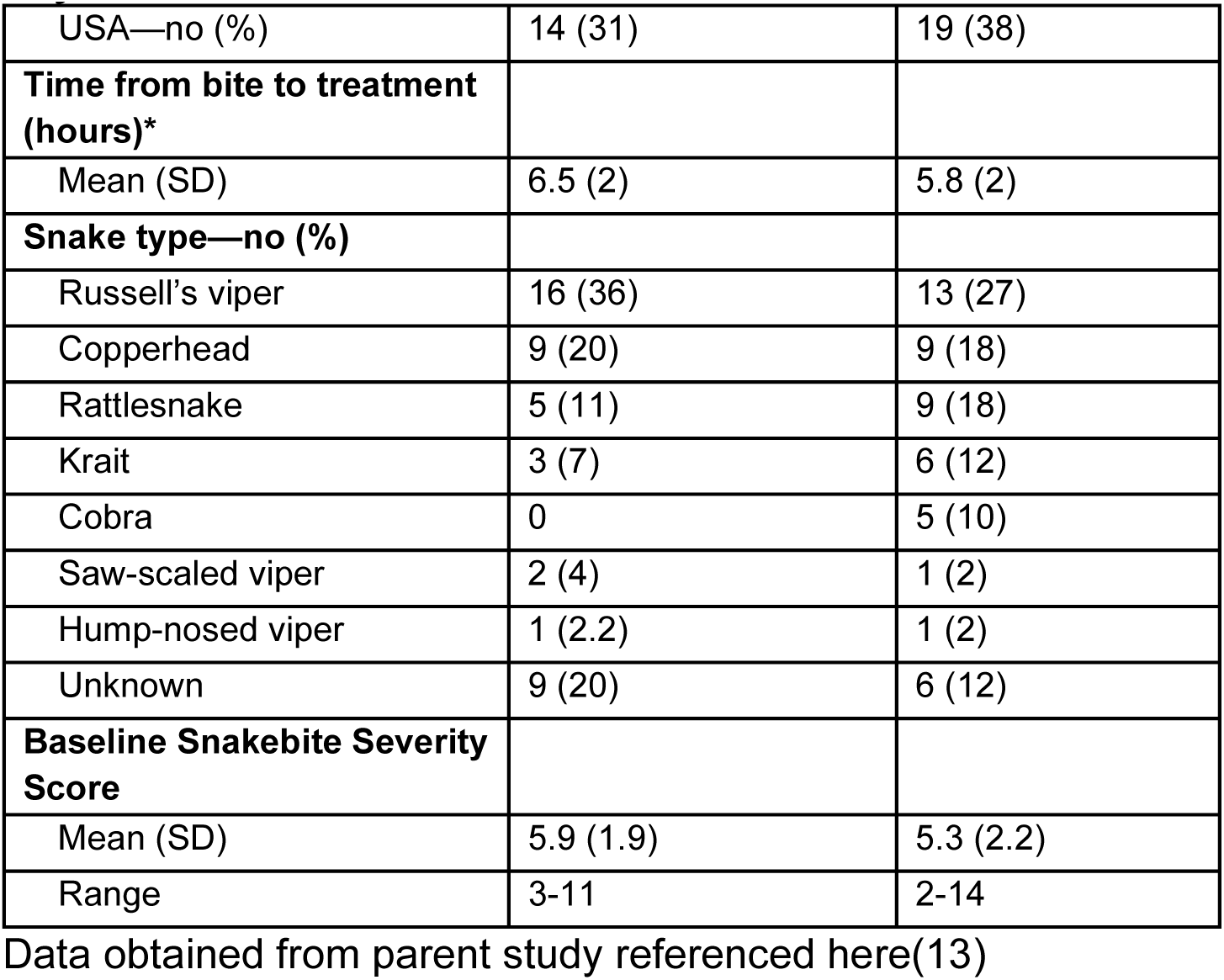
Participants Characteristics (N=95)

| Characteristic | Varespladib (n=45) | Placebo (n=50) |
| --- | --- | --- |
| **Age—year** |  |  |
| Mean (SD) | 38 (19) | 42 (16) |
| 5-10 years—no (%) | 1 (2) | 2 (4) |
| 11-17 years—no (%) | 5 (11) | 2 (4) |
| 18 years and older—no (%) | 39 (87) | 46 (92) |
| **Sex** |  |  |
| Male—no (%) | 35 (78) | 39 (78) |
| Female—no (%) | 10 (22) | 11 (22) |
| **Country** |  |  |
| India—no (%) | 31 (69) | 31 (62) |
| USA—no (%) | 14 (31) | 19 (38) |
| **Time from bite to treatment (hours)\*** |  |  |
| Mean (SD) | 6.5 (2) | 5.8 (2) |
| **Snake type—no (%)** |  |  |
| Russell's viper | 16 (36) | 13 (27) |
| Copperhead | 9 (20) | 9 (18) |
| Rattlesnake | 5 (11) | 9 (18) |
| Krait | 3 (7) | 6 (12) |
| Cobra | 0 | 5 (10) |
| Saw-scaled viper | 2 (4) | 1 (2) |
| Hump-nosed viper | 1 (2.2) | 1 (2) |
| Unknown | 9 (20) | 6 (12) |
| **Baseline Snakebite Severity Score** |  |  |
| Mean (SD) | 5.9 (1.9) | 5.3 (2.2) |
| Range | 3-11 | 2-14 |

Data obtained from parent study referenced here(13)

At baseline, the highest median SSS scores were observed in the local wound (2 IQR [2, 2]), hematological (1 [0, 3]), and nervous (1 [0, 1]) subscores (Table 2). Over time, a general decline in symptom burden was noted across all subscores, with marked reductions by day 14. By day 28, median scores were lowest across all domains, particularly in the gastrointestinal and cardiovascular systems both at 0 (IQR 0, 0).

**Table 2:**
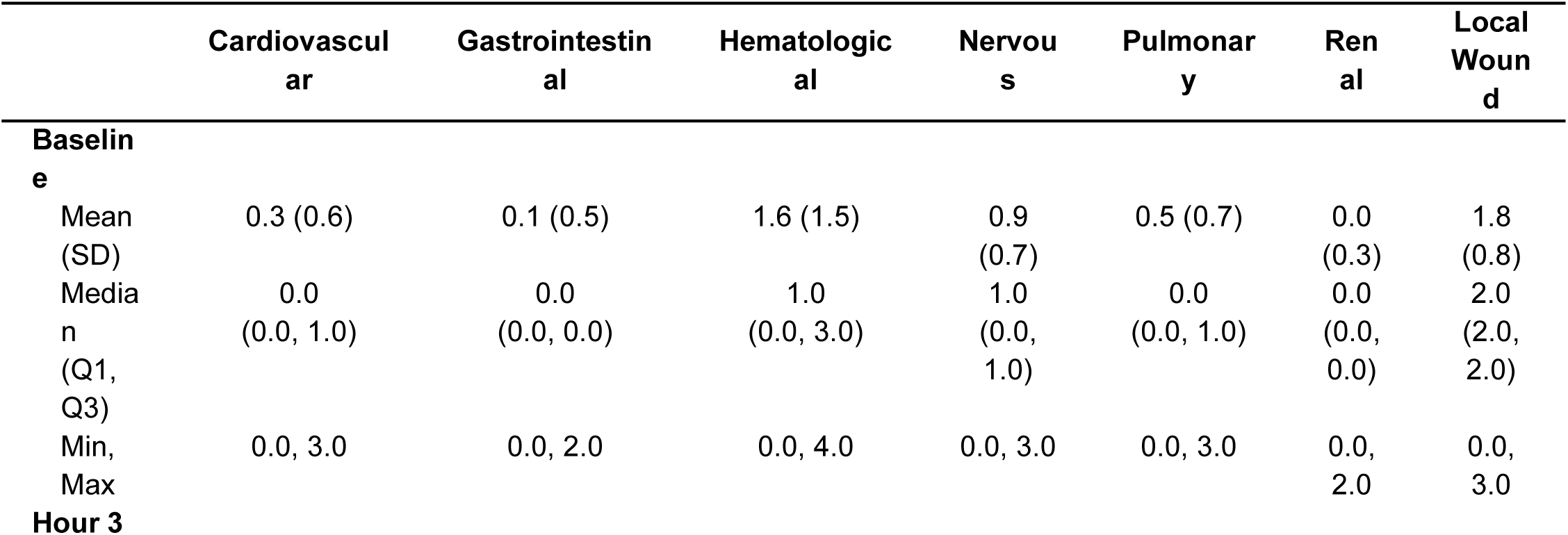
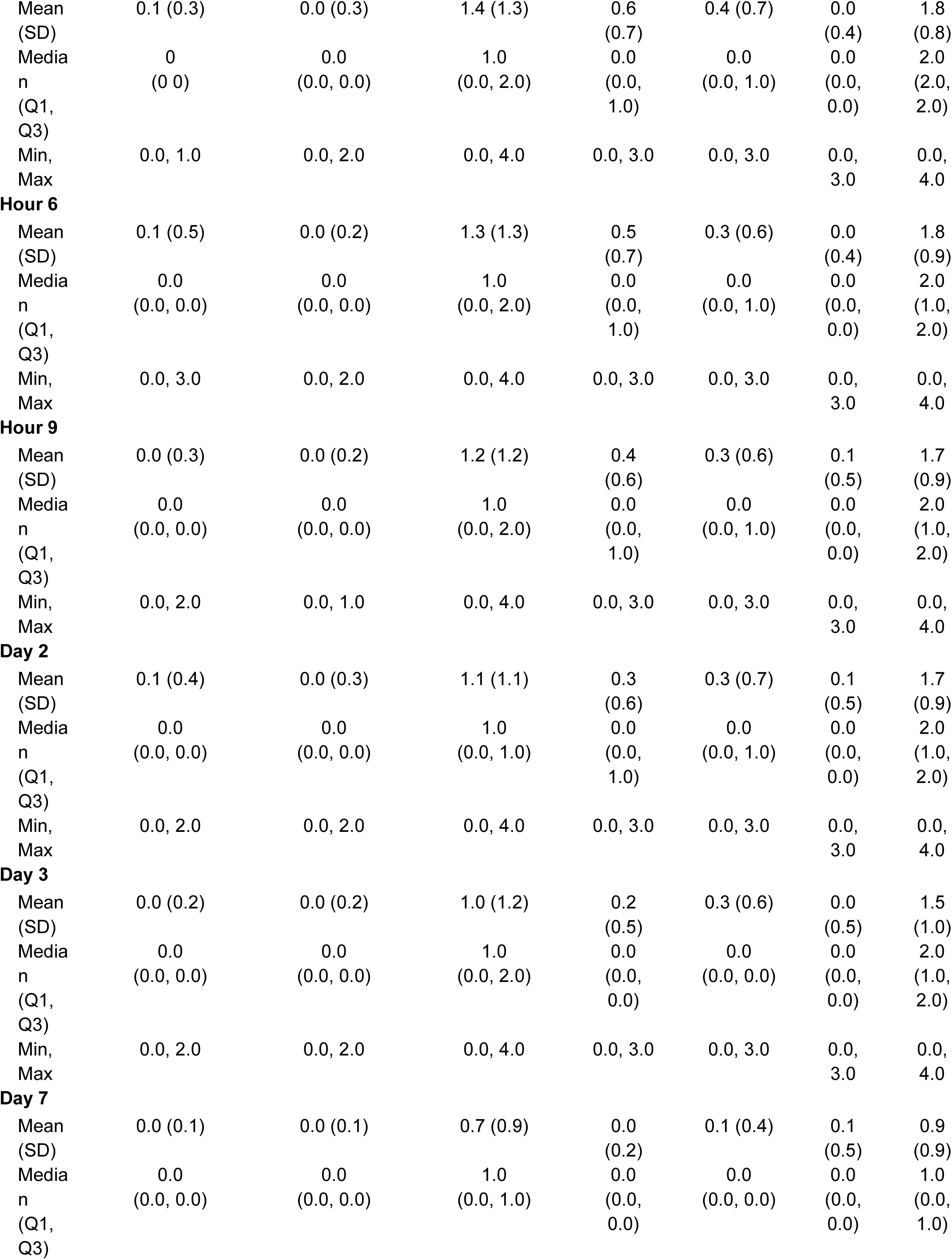
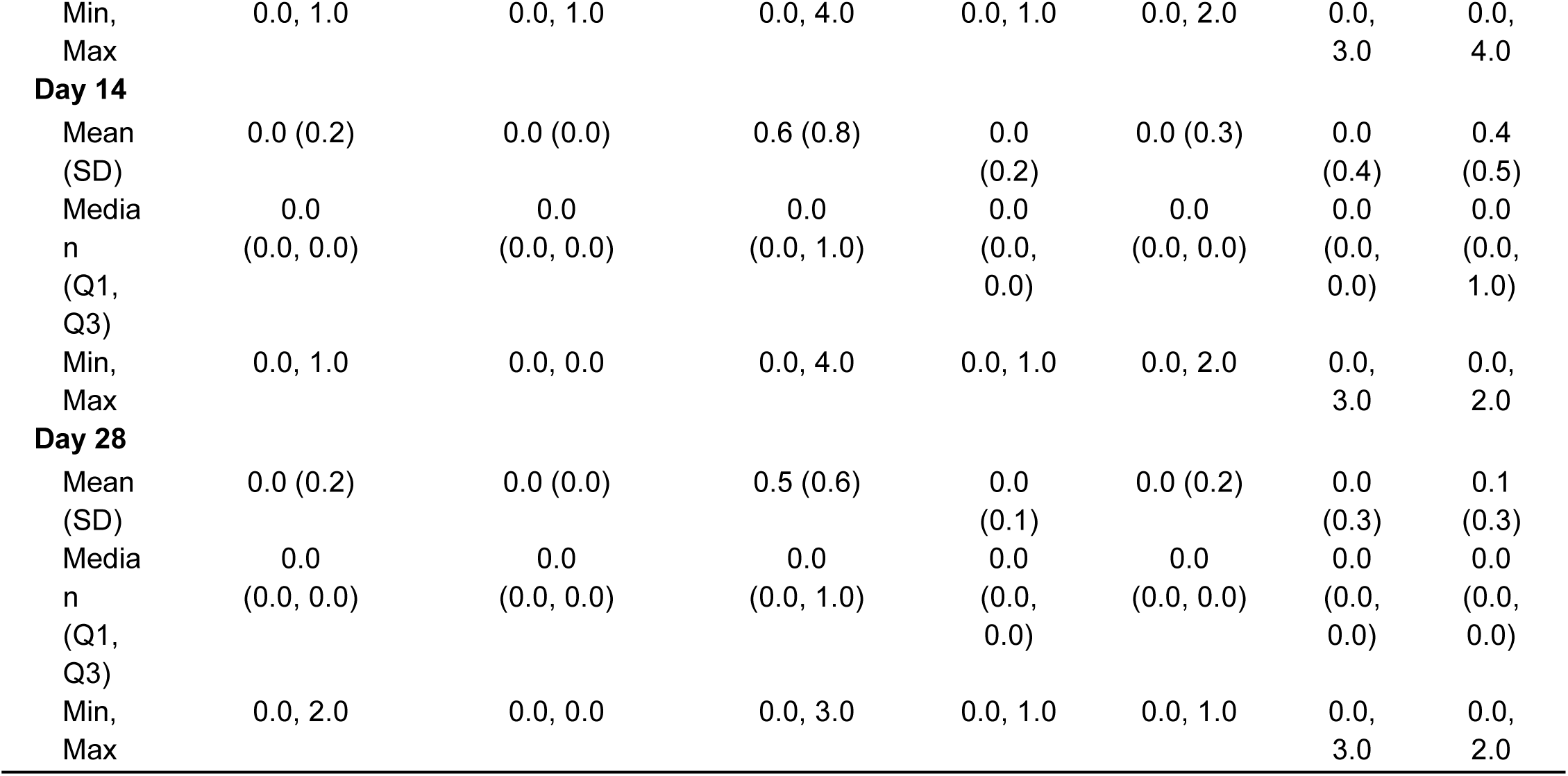
Overall Descriptive statistics for the SSS Subscale across time points (n=95)

We explored the SSS using several summary approaches with different time points and summary measures (Mean SSS between different time points; raw SSS score; SSS differences; and AUC) (Table 3). As expected, given the inclusion of additional non-negative symptom domains, summary measures derived from the 6-item SSS were higher than those from the 3-item SSS. This was particularly true for AUC and difference-based metrics. For the 3-item SSS, the median score was 2.5 (IQR 1.5, 3.0) for days 3 and 7 combined, 2.0 (1.0, 3.0) at day 7, and declined to 1.5 (0.5, 2.0) for days 7 and 14 combined. For the 6-item SSS, the corresponding median summed scores were 2.5 (1.5, 3.5), 2.0 (1.0, 3.0), and 1.5 (0.8, 2.5), respectively.

**Table 3:**
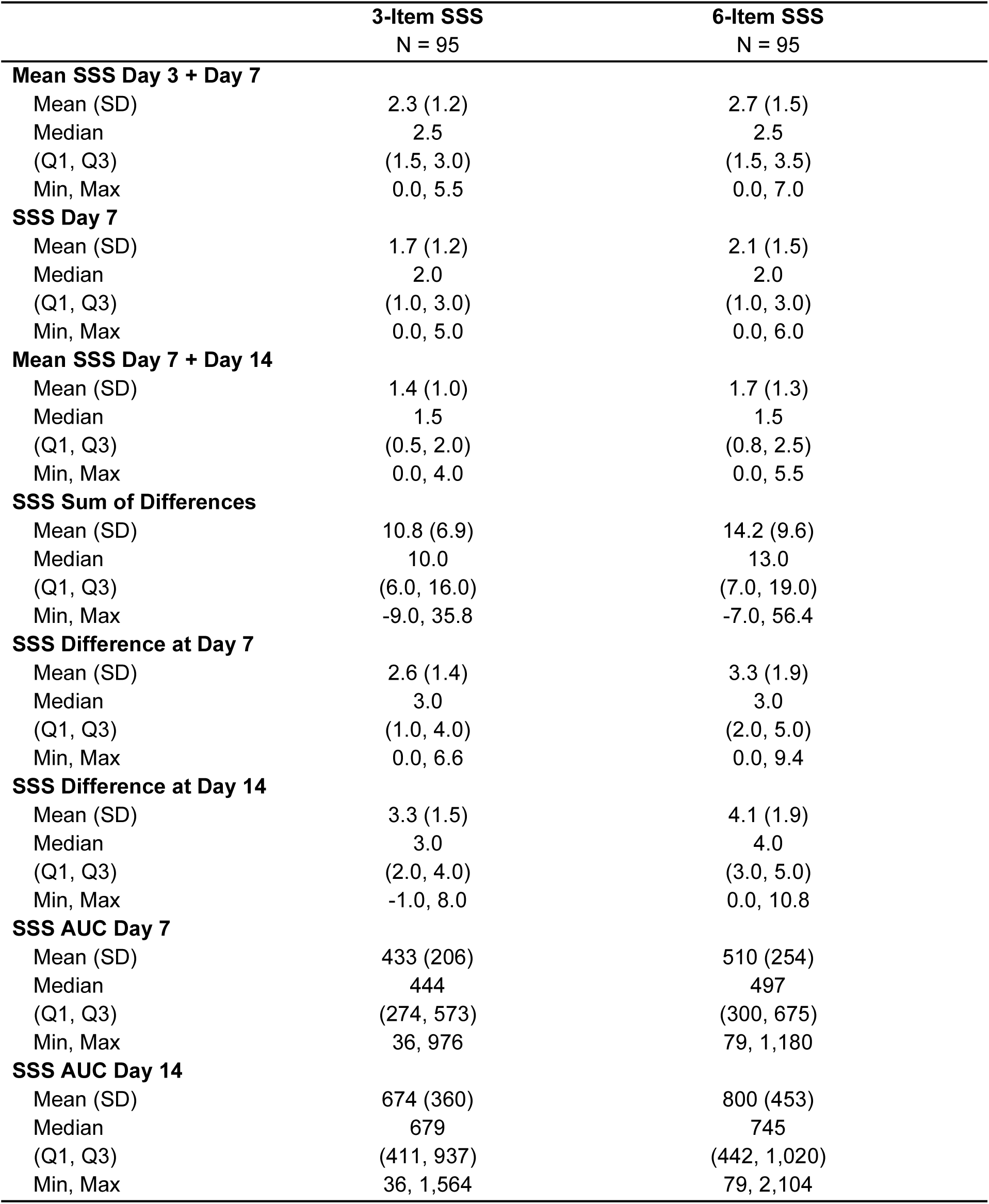
Descriptive Statistics for the 3-Item and 6 item SSS at different Timepoints.

For symptom burden over time, AUC medians were higher for the 6-item SSS at day 7 (497 [300, 675] vs 444 [274, 573)] and day 14 [745 (442–1,020) vs 679 (411–937)]. Difference-based summaries also trended higher for the 6-item scale (sum of differences 13 [7–19] vs 10 [6–16]; day 14 difference 4 [3–5] vs 3 [2–4]).

### Evidence of reliability

For evidence of reliability, we observed that the SSS items had different responsiveness over time (Table 2). The local wound and hematological subscores were stable until day 3, then improved consistently through day 28 with notable persistence in hematologic scores of 1 in a substantial portion of the patients. These subscores showed no indication of floor or ceiling effect at baseline. The nervous system and pulmonary subscores showed a rapid reduction from baseline to the early follow-up times.

While no floor effect was observed at baseline, there was an increasing sign of floor effect at follow-up as is expected with patient recovery. Cardiovascular, renal and gastrointestinal subscores had small variations over time, with signs of floor effect at baseline and follow-up. This illustrates that the SSS is a multi-dimensional composite endpoint that provides evidence of the response of each clinical venom effect (subscore) over disease progression and recovery. The different timelines for the resolution of some subscores highlight the need for an SSS application that integrates the variation in scores over time.

The intraclass correlation coefficients (ICCs) assessing agreement across repeated measures for the 3- and 6-item SSS at day 7 and day 14 were 0.7 (95% CI: 0.6-0.8) and 0.8 (95% CI: 0.6-0.8), respectively. Both estimates indicate good reliability, with the 6-item version showing slightly higher inter-rater agreement. The associated F-values were statistically significant (p < 0.01), confirming consistent ratings across raters. Full results are shown in **Table 4**

**Table 4:**
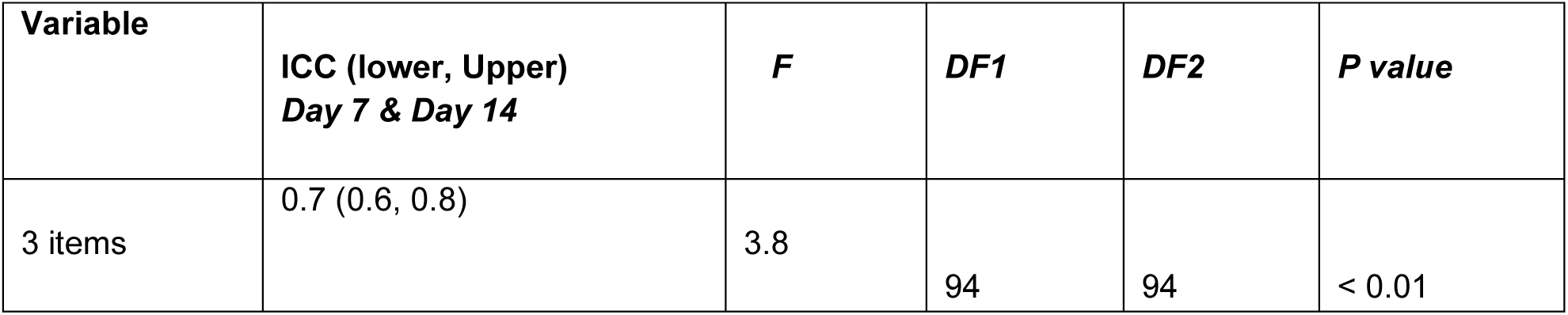
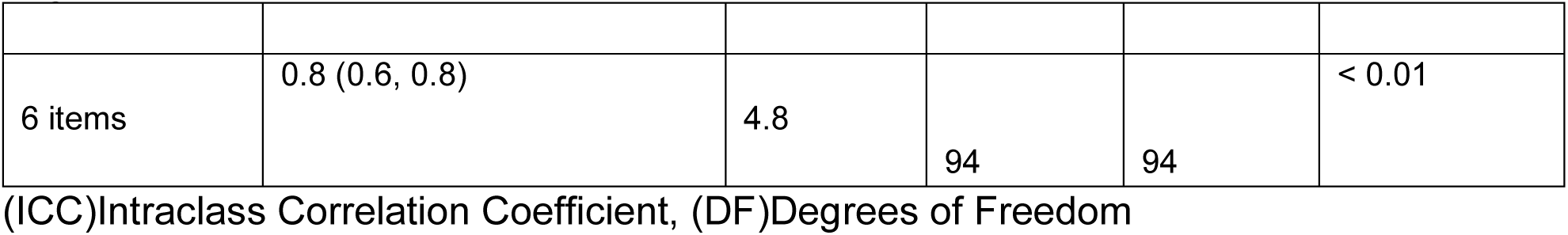
Average fixed raters ICC comparison between 3- and 6-item SSS at day 7 and 14:

| Variable | ICC (lower, Upper)<br>Day 7 & Day 14 | F | DF1 | DF2 | P value |
| --- | --- | --- | --- | --- | --- |
| 3 items | 0.7 (0.6, 0.8) | 3.8 | 94 | 94 | < 0.01 |
| 6 items | 0.8 (0.6, 0.8) | 4.8 | 94 | 94 | < 0.01 |

(ICC)Intraclass Correlation Coefficient, (DF)Degrees of Freedom

#### Evidence of Validly related to internal structure

The principal component analysis (PCA) identified three main components explaining variance across the SSS subscores. Pulmonary, nervous and local wound subscores loaded strongly on PC1 (0.8, 0.7, and −0.7, respectively), while hematological symptoms were highly associated with PC2 (loading = 0.9). Cardiovascular and gastrointestinal subscores showed higher loadings on PC3 (0.6 and 0.6, respectively). Communality values ranged from 0.3 (renal) to 0.8 (hematological), indicating varying levels of shared variance across subscores. Detailed loadings and component contributions are presented in **Table 5**.

**Table 5:** PCA Loading Table for All SSS subscores and patients: Three Principal Components.

| Subscore | PC1 | PC2 | PC3 | Communality | Uniqueness | Complexity |
| --- | --- | --- | --- | --- | --- | --- |
| Cardiovascular | 0.2 | 0.2 | 0.6 | 0.5 | 0.4 | 1.0 |
| Gastrointestinal | -0.0 | -0.0 | 0.6 | 0.4 | 0.5 | 1.0 |
| Hematological | -0.0 | 0.9 | -0.0 | 0.8 | 0.1 | 1.0 |
| Local Wound | -0.7 | 0.0 | 0.0 | 0.5 | 0.4 | 1.0 |
| Nervous | 0.7 | -0.4 | -0.0 | 0.7 | 0.2 | 1.7 |
| Pulmonary | 0.8 | 0.1 | 0.0 | 0.6 | 0.3 | 1.0 |
| Renal | 0.1 | 0.2 | -0.5 | 0.3 | 0.6 | 1.8 |

The biplot of principal component analysis (PCA) in **Figure 1** displays the loadings of each SSS subscore on the first two principal components (PC1 and PC2). Pulmonary and nervous subscores load strongly on PC1, indicating a high contribution to this component, while the hematological subscore is most aligned with PC2. Local wound subscore loads negatively on PC1, showing distinct clustering. Renal and cardiovascular subscores contribute modestly to both PC1 and PC2, whereas gastrointestinal shows minimal loading on either. This visual representation complements the PCA loading **Table 5** above and highlights the differential contributions of subscores to underlying symptom structures

**Figure 1:**
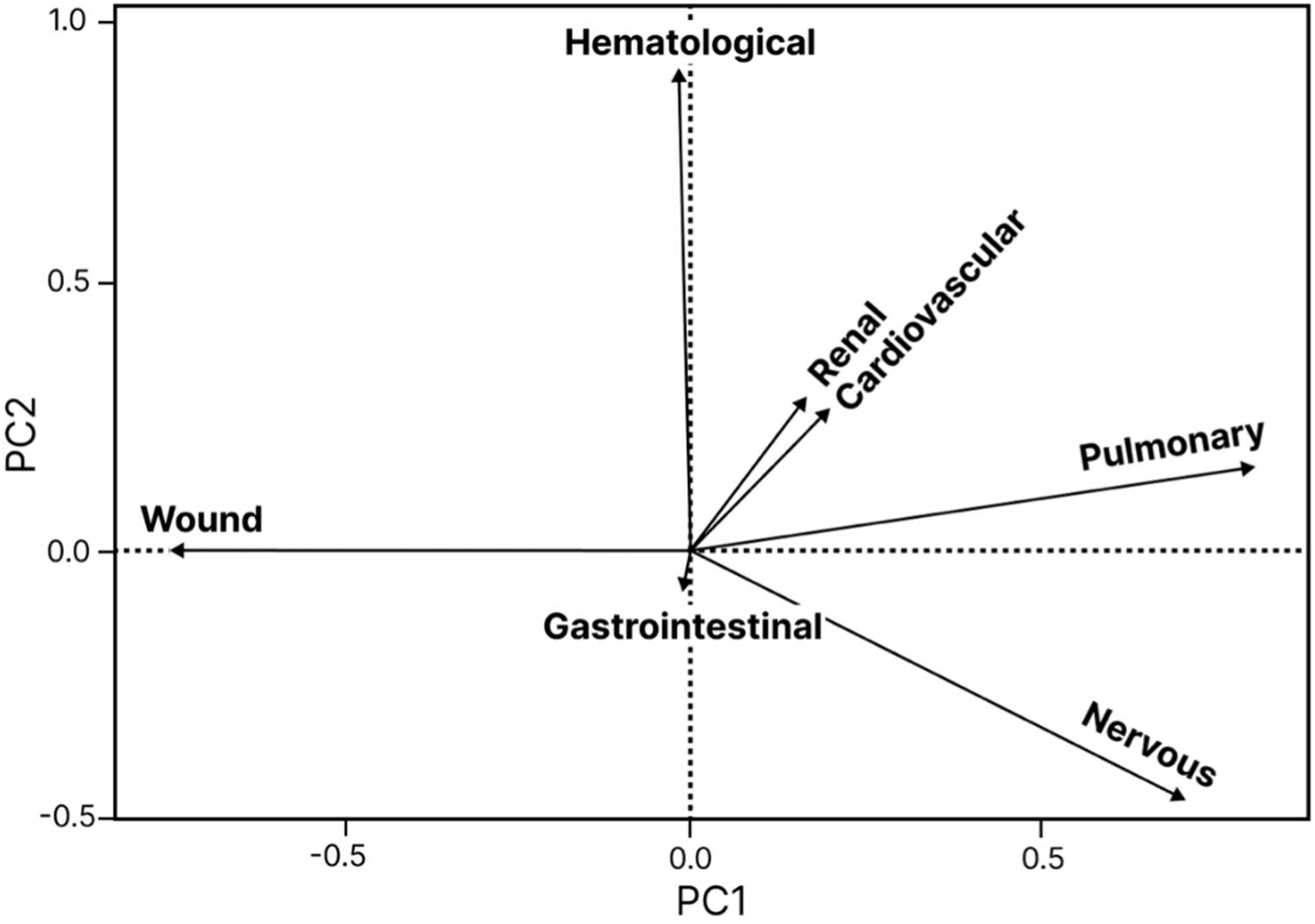
PCA Loadings plot for PC1 and PC2, All SSS subscores and patients.

The scree plot (left) and cumulative variance plot (right) in **Figure 2** display the variance explained by each principal component derived from the SSS subscores. The first three principal components explain a cumulative 62% of the total variance (26% by PC1, 19% by PC2, and 17% by PC3), suggesting these three components capture most of the data structure. By the sixth component, 100% of the variance is explained, indicating all components together account for the full variability across subscores

**Figure 2:**
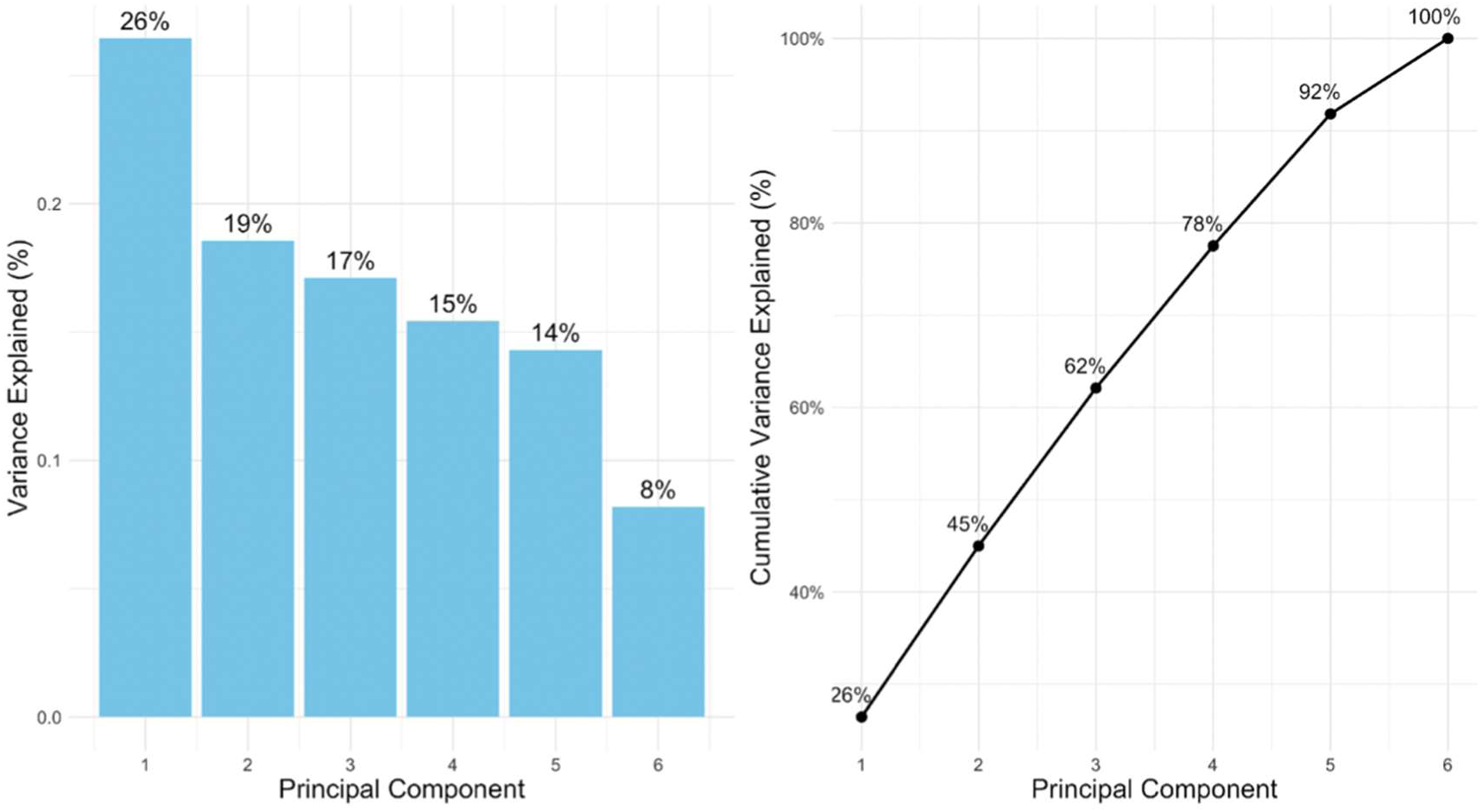
PCA Scree plot: All SSS subscores.

#### Evidence of validity related to associated variables

We observed a strong positive correlation among the alternative SSS applications that capture the SSS variation over time (e.g., AUC Day 7 and Day 14, mean scores, and sum/difference scores) and the absolute SSS at 7 days, with correlation coefficients ranging from 0.7 to 0.9. Conversely, the SSS applications that use differences from baseline (Sum of Differences or Absolute Differences) demonstrated low correlation with the time-varying measures of the SSS (Figure 3).

**Figure 3:**
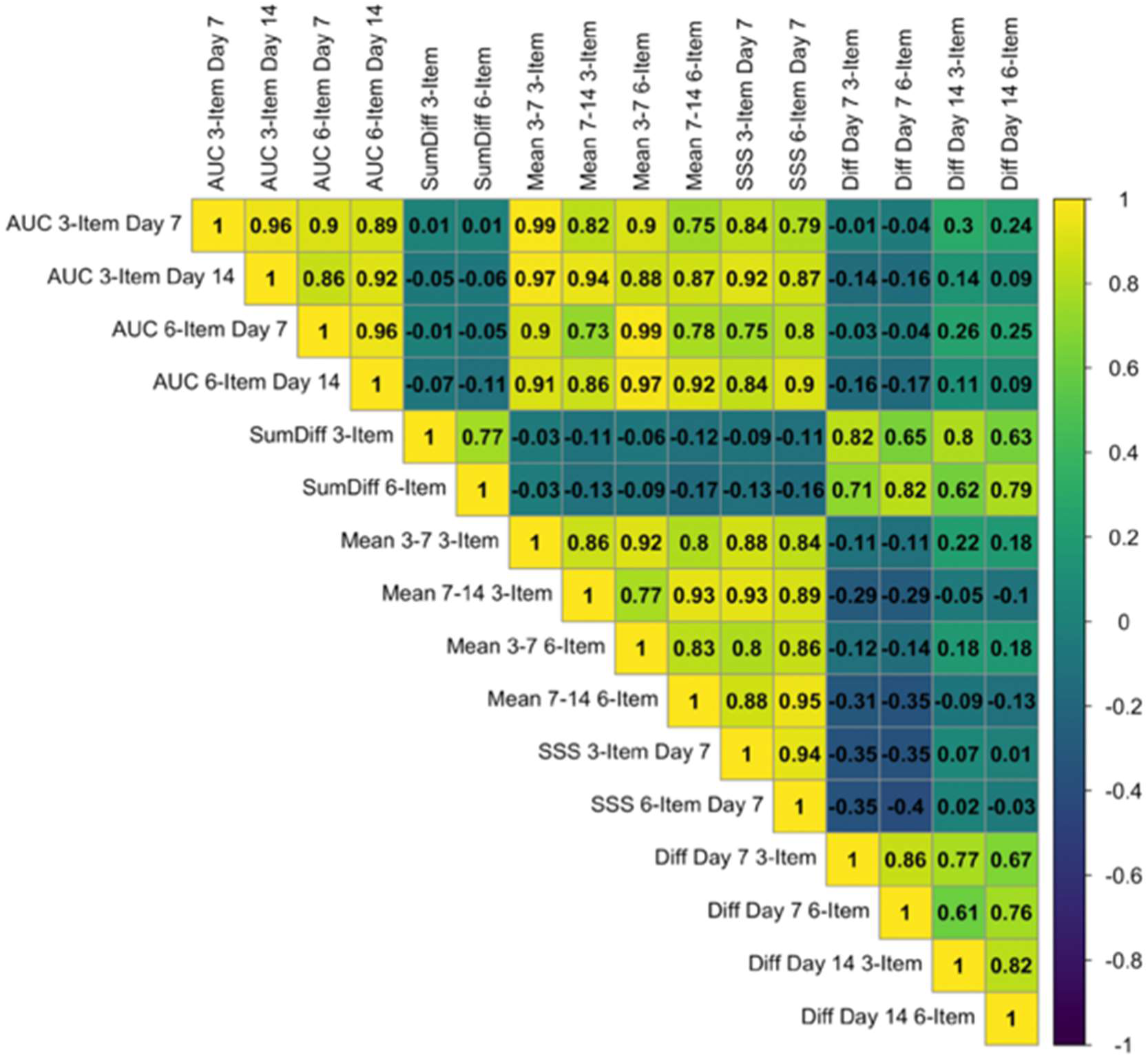
Correlation among different SSS applications (3- and 6-item).

A similar pattern was observed when associating the SSS applications with other patient-reported outcome measures, for both the 3- and 6-items SSS. Moderate positive correlations were observed between SSS applications that capture the SSS variation over time and PGIC (Patient Global Impression of Change), NPRS (Numeric Pain Rating Scale), and CGI-I (Clinical Global Impression-Improvement), especially at day 14 (e.g., PGIC and mean 7-14: *r*= 0.4; NPRS and AUC day 14: *r* = 0.4). Correlations with the PSFS were also positive; to facilitate comparison, the direction was inverted (original r = −0.4; reported as r = 0.4), as higher PSFS scores indicate lower patient functionality. The SSS applications that use differences from baseline (Sum of Differences or Absolute Differences) demonstrated low correlation with other patient-reported outcomes (Figure 4A and B).

**Figure 4.**
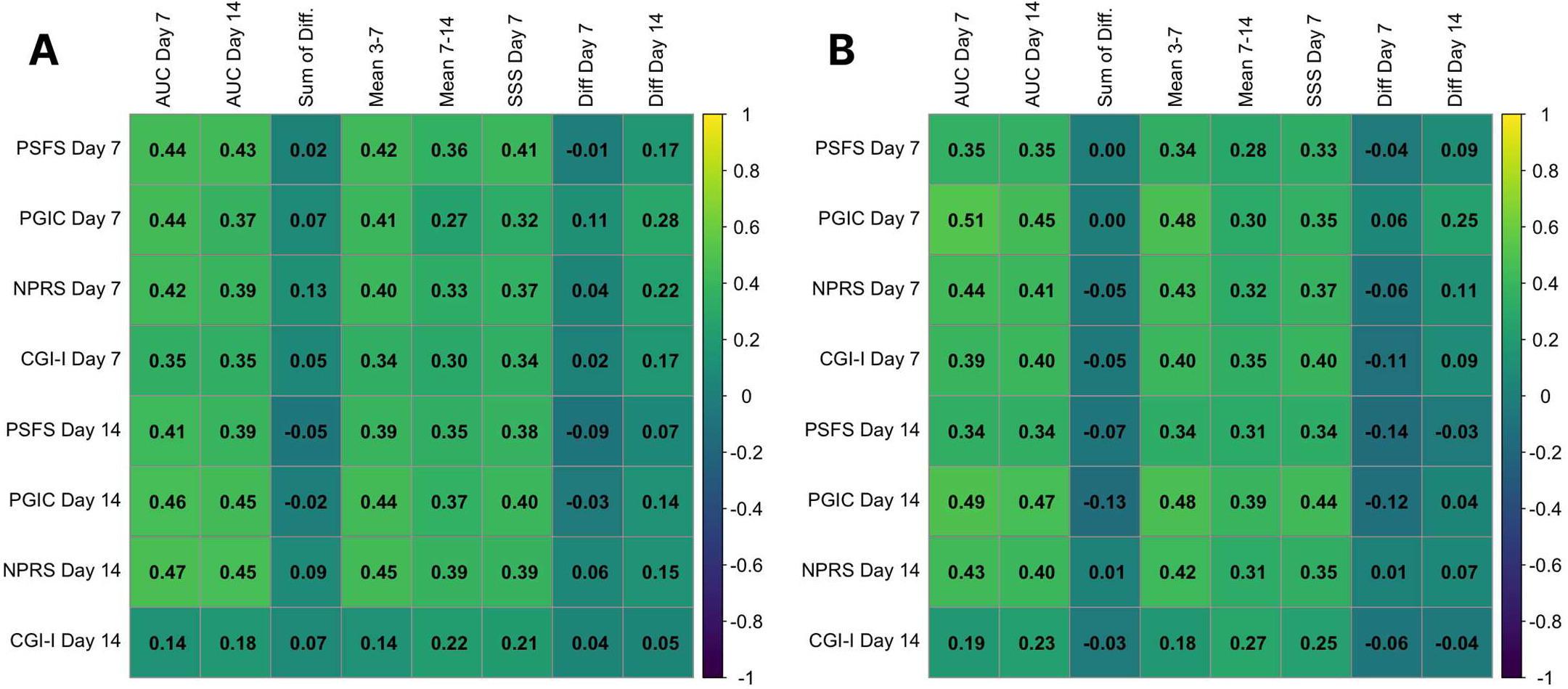
A and B: Correlation among SSS alternative scoring methods (3-item, A, 6-item B) and other patient outcome metric outcomes. To facilitate comparison, the direction of the PSFS correlation was inverted (original r = −0.44 and −0.35; reported as r = 0.44 and 0.35), as higher PSFS scores indicate lower patient functionality.

## DISCUSSION

To our knowledge, this study is the first comprehensive psychometric evaluation of the SSS using the American Psychological Association Standards, as well as multiple potential SSS applications that characterize disease courses in SBE. We evaluated reliability by describing SSS scores over time, depicting ceiling and floor effects, and evaluating temporal stability. Internal structure was evaluated using subscore correlations and PCA. External (convergent) validity was evaluated through the correlation of multiple SSS applications with each other and with additional SBE-related outcomes. Our key findings from this research describe the reliability and validity of the SSS as well as several SSS applications that characterize disease courses in SBE.

The SSS demonstrated good reliability without baseline ceiling or floor effects and with good temporal stability. For evidence of reliability, we observed that the SSS items had different responsiveness over time. This illustrates that the SSS is a multi-dimensional composite endpoint that provides evidence of the response of each clinical venom effect (subscore) over disease progression and recovery. The different timelines for the resolution of some subscores highlight the need for an SSS application that integrates the variation in scores over time. Although the SSS was designed as a research tool, it has been applied clinically. The multi-dimensionality of SSS subscores should be considered if making decisions regarding diagnosis and treatment to either prevent costly administration of unnecessary treatment(26) or prevent failure to administer appropriate evidence-based treatments (12). Analysis of the interclass correlation between assessment scores at day 7 and day 14 produced high correlation and supported high internal stability(13).

The evaluation of the SSS internal structure confirmed the multi-dimensionality of the SSS subscores that independently capture unique responses of SBE clinical venom effects. When analyzing the SSS subscores at baseline, we observed that the subscores do not have a high correlation. This is expected as SBE symptoms are multi-dimensional, particularly across global snake species. This result illustrates the unique contribution of each subscore to the assessment of SBE severity. We confirmed this multi-dimensionality through PCA analysis. The models with two or three components explain a substantial proportion of the variance. We observed that local wound, nervous system, and hematological subscore (3-item SSS) do not lie on the same principal components, consistent with unique contributions of each subscore.

The SSS applications that captured variation over time (AUC and Mean SSS) had low correlation with those that captured differences from baselines (Sum of the Differences and Absolute Differences from baselines). This result is expected as both types of SSS applications capture different responsive behaviors of the SSS. When anchored to external additional SBE related outcomes, the SSS applications that capture variation over time (Area Under the Curve and Mean Over Time SSS) showed better external validity compared to methods capturing differences from baseline (Sum of the differences and subscore -level differences). Hence, the SSS applications that capture variation over time have better correlation with pain, functionality, and perception of improvement. In summary, the SSS demonstrates adequate psychometric properties of reliability and external validity. Its internal structure confirms its design as a multi-dimensional composite endpoint. The SSS applications that integrate variation over time are preferred.

Despite its widespread use, the SSS has some limitations from a regulatory perspective. As a single unweighted composite endpoint, it aggregates clinically heterogeneous subscores—including local tissue injury and systemic toxicity—into a total score that may obscure differential treatment effects across biologically distinct processes. This structure creates a classic composite endpoint problem, in which changes in less clinically meaningful or more prevalent components may dominate the total score, while effects on rarer but more serious outcomes are diluted. One potential modification to improve regulatory alignment would be to explicitly separate local wound effects from systemic toxicity into distinct domains, while retaining a prespecified total score that applies differential weighting to these components based on clinical importance and expected treatment mechanism.

We would like to acknowledge the limitations presented in this study. First, the data used for this study was collected as secondary analysis and was not intended for this purpose. This may reasonably limit the representativeness of our findings. Second, this study relies on self-report data which may be impacted by social desirability bias, misunderstanding of questions, or inconsistent responses. Third, due to the nature of the SSS, principal component analyses conducted on the SSS subscores use ordinal data, not continuous data. Finally, venom heterogeneity, bite location, and individual differences in physiological response can impact symptomatic presentation. It is possible that in a larger sample, and with greater representation of different venom types, we might see increased variation in scores across SSS subscales.

## CONCLUSION

The implications of this research include the understanding that the SSS demonstrates adequate psychometric properties of reliability and external validity. Its internal structure confirms its design as a multi-dimensional composite endpoint. The SSS applications that integrate variation over time are preferred. Our findings support the reliability and validity of the SSS as a tool to assess SBE symptomatic severity. Further research is needed to understand how type of venom, bite location, and patient characteristics influence SSS subscores.

## Data Availability

Data available upon request to authors.

## Acknowledgements

We acknowledge the efforts of the many investigative teams and partners managing the protocol and the staff itself beyond those listed here.

